# A Phase 1 Study of Novel Antiplatelet Agent to Overcome Pharmacogenomic Limitations of Clopidogrel

**DOI:** 10.1101/2024.03.21.24304696

**Authors:** Anil Pareek, Nitin Chandurkar, Vivek Raut, Kumar Naidu

## Abstract

**Background:** Clopidogrel is the most commonly prescribed thienopyridine as part of dual antiplatelet therapy for the treatment of cardiovascular diseases. However, Clopidogrel responsiveness shows variability based on CYP2C19 polymorphism. Therefore, we planned a study with an objective of evaluating safety, tolerability, pharmacodynamics and pharmacokinetics of a novel thienopyridine antiplatelet agent AT-10 in healthy Indian subjects in comparison with standard dosage regimen of Clopidogrel based on their CYP2C19 genotyping.

**Methods:** Two CYP2C19 genotype based groups were identified i.e., Poor Metabolizer and Extensive Metabolizers with 20 subjects in each group (N=40) for participating in a randomized, two period, cross-over study. Each study period lasted 6 days including administration of loading and maintenance doses of AT-10 (40mg/10mg) or Clopidogrel (300mg/75mg). The pharmacokinetics and pharmacodynamics were assessed on day 1 and day 6 at several time intervals.

**Results:** Overall result of pharmacodynamic parameters showed that, mean % Inhibition of Platelet Aggregation between AT-10 and Clopidogrel in all subjects at Post-dose 6hr (loading dose) (AT-10: Clopidogrel; 73.30% vs. 18.53%) and Post-dose 6 hr on Day 6 (maintenance dose) (AT-10: Clopidogrel; 83.41% vs. 51.19 %) obtained from the AT-10 group was significantly higher than the Clopidogrel group. Further, %inhibition of platelet aggregation from AT-10 treatment in poor metabolizer group was significantly higher than the Clopidogrel treatments in extensive metabolizer group.

Overall pharmacokinetic comparison in all subjects indicates that AT-10 gives greater exposure to active Metabolite MP-H4 than Clopidogrel.

**Conclusion:** AT-10 may emerge as a promising anti-platelet drug and can be further developed in clinical studies for the unmet medical needs of cardiovascular diseases. CTRI Registration: CTRI/2021/03/032206

**Clinical Perspectives:**

- AT-10 (2-oxo-Clopidogrel) is a novel thienopyridine P2Y12 inhibitor under clinical development in India.
- It is a Clopidogrel derivative that is metabolically converted to produce the active thiol metabolite similar to Clopidogrel.
- The response to AT-10 was found unaffected by genetic CYP2C19 polymorphisms to the extent of its influence in Clopidogrel, thus eliminating the need for characterization of Clopidogrel responsiveness and thereby maintaining effectiveness in all patients, including those patients who are identified as CYP2C19 poor and intermediate metabolizers.
- Further studies are required to investigate the influence of CYP2C19 functional polymorphisms on the response to AT-10 dose in diverse clinical settings, and especially on the risk of recurrent thrombotic events during AT-10 treatment as compared to Clopidogrel therapy.

## INTRODUCTION

Cardiovascular diseases remain the leading cause of mortality worldwide. Estimated 20.5 million deaths were reported globally due to cardiac conditions in 2021 which showed a marked increase from 12.1 million CVD deaths recorded in 1990^1^. Of these deaths, more than 50% are secondary to coronary artery disease, with acute coronary syndrome (ACS) accounting for approximately 10% of all admissions presenting to emergency care physicians^2^. Platelets play a key role in the development of ACS, as plaque rupture is followed by platelet adhesion, activation, and aggregation, leading to thrombosis formation^3^. Antiplatelet drugs serve as a first-line antithrombotic therapy for the management of acute ischemic events and the prevention of secondary complications in vascular diseases. Numerous antiplatelet therapies have been developed; however, currently available agents are still associated with inadequate efficacy, risk of bleeding, and variability in individual response^4^. Dual Antiplatelet therapy with Aspirin and P2Y12 inhibitors, drugs from thienopyridine class (Ticlopidine, Clopidogrel, and Prasugrel) is therefore the cornerstone of medical management in patients with ACS, those undergoing percutaneous coronary Intervention (PCI), prevention of stent thrombosis and is necessary in both the acute phase and long-term maintenance therapy^5–7^. Despite these benefits, many patients continue to have recurrent atherothrombotic events while receiving standard dual antiplatelet therapy^8^. Amongst all P2Y12 inhibitors, Clopidogrel is the most commonly prescribed thienopyridine with acceptable safety profile. However, the pharmacodynamic response to Clopidogrel varies widely from subject to subject, and about 25% of patients treated with standard Clopidogrel doses display low *ex vivo* inhibition of ADP-induced platelet aggregation^9^. While the use of Clopidogrel has undoubtedly been a beneficial advance in the treatment of ACS with regard to both short and long term morbidity and mortality^10, 11^, it remains a treatment with several shortcomings.

Clopidogrel is a prodrug and its clinical efficacy appears to be a function of the amount of enzymatically derived active thiol metabolite formed. Clopidogrel is first metabolized to the intermediate metabolite, 2-oxo-Clopidogrel, followed by metabolism to an active thiol metabolite in vivo. Notably, both metabolic steps leading to thiol formation have been shown to be predominantly dependent on CYP2C19 and, to a lesser extent, CYP3A4. As a prodrug, around 85% of Clopidogrel *in vivo* is inactivated by carboxylesterase-1 (CES-1) and the remaining 15% is converted to the active metabolite H4 via a two-step process in liver, which is predominantly influenced by cytochrome CYP2C19 polymorphisms. Various CYP enzymes are involved for converting Clopidogrel to its active metabolite (H4). First metabolic step is predominantly dependent on CYP2C19 (45%) and second step on CYP3A4 enzyme (40%). However, involvement of CYP2C19 during second step is only 21%. Since CYP2C19 is involved in both metabolic steps, any changes in its activity will have significant impacts on the formation of the active thiol metabolite and hence, on the response to treatment^12,13^. The metabolism of Clopidogrel to its active metabolite can be impaired by genetic variations in CYP2C19 or by drugs that inhibit CYP2C19, such as Omeprazole or Esomeprazole.

It has been established that not all of the patients receiving Clopidogrel benefit to the same extent, and it has been shown that patients with a Poor Metabolizers (PM) genotype status for CYP2C19 are at increased risk of ischemic events after percutaneous coronary intervention ^5,14^ suggesting that CYP2C19 polymorphism is a clinically relevant determinant of response to Clopidogrel. In 2010, the USFDA had imposed a box warning on Clopidogrel bisulfate tablets (Plavix) stating that “Effectiveness of Clopidogrel bisulfate depends on conversion to an active metabolite by the cytochrome P450 (CYP) system, principally CYP2C19 enzyme. Consider use of another platelet P2Y12 inhibitor in patients identified as CYP2C19 poor metabolizers” ^15^. It means there is a well-established relationship exists between CYP2C19 deficient pharmacogenomic populations and Clopidogrel drug response that result in diminished effectiveness in PMs because of impaired metabolism of Clopidogrel.

Patients with carriers of reduced-function CYP2C19 alleles (poor and intermediate metabolizers) will produce lower levels of Clopidogrel’s active thiol metabolite. Due to which such patients shows diminished platelet inhibition, and are at higher risk of major adverse cardiovascular events, including a threefold greater risk of stent thrombosis ^16,17,18^ hence newer antiplatelet are preferred in such patients. Although, the newer agents like Prasugrel and Ticagrelor are devoid of such problems (genetic polymorphism) the CV benefits comes with cost of increased bleeding risk that includes fatal intracranial hemorrhage^19^.

There is a considerable heterogeneity observed in the activity of CYP450 enzymes in humans and it has become apparent that individuals have polymorphisms in CYP2C19 resulting in low or non responsiveness to Clopidogrel known as ‘Clopidogrel resistance’ ^16,20^. The frequency of CYP2C19*2 allele associated with PM type reported to be 47.23% in Coronary Artery Disease (CAD) patients in general population in India^21^. Studies conducted by Adithan et. al^22^ have reported incidence of CYP2C19 polymorphism of around 37.9% in south Indian general population and 35.5% by Kavita et al^23^ in western Indian general population.

In the recently published real-world data composing East Asians who had undergone Drug Eluting Stent (DES) implantation and received Clopidogrel based antiplatelet therapy indicated that, Intermediate Metabolizers (IM) or PMs (PM) which was close to 62.1% of the total population evaluated had a higher risk of cardiac death, myocardial infarction, and stent thrombosis at 5-year follow-up compared with normal metabolizers^24^. Further, authors have also reported that the risk of cardiac death, myocardial infarction and stent thrombosis in IMs and PMs was much higher within 1 year after DES implantation. The most common CYP2C19 loss of function allele is *2 with allele frequencies of ∼15% in Caucasians and Africans, and 29–35% in Asians. These data indicates that the prevalence of IM and PM is more in Asian population than that of the western population ^18,24^. This high prevalence of CYP2C19 polymorphism renders Clopidogrel ineffective or adds variability in effectiveness in poor and intermediate metabolizers respectively.

Hence, to overcome the shortcomings of Clopidogrel associated mainly with CYP2C19 metabolism, we have developed a novel antiplatelet agent AT-10 (2-oxo-clopidogrel), which is a Clopidogrel derivative that is metabolically converted in one CYP-dependent step, to produce the active metabolite similar to the approved listed drug Plavix (Clopidogrel bisulfate tablet). A major advantage of the prodrug AT-10 is its more efficient metabolism through one CYP-dependent step, as compared to the two-step process for Clopidogrel involving several CYP450 isoenzymes. The metabolic pathway of Clopidogrel and AT-10 (2-oxo-clopidogrel) is depicted in **Figure 1**.

**Fig.1:**
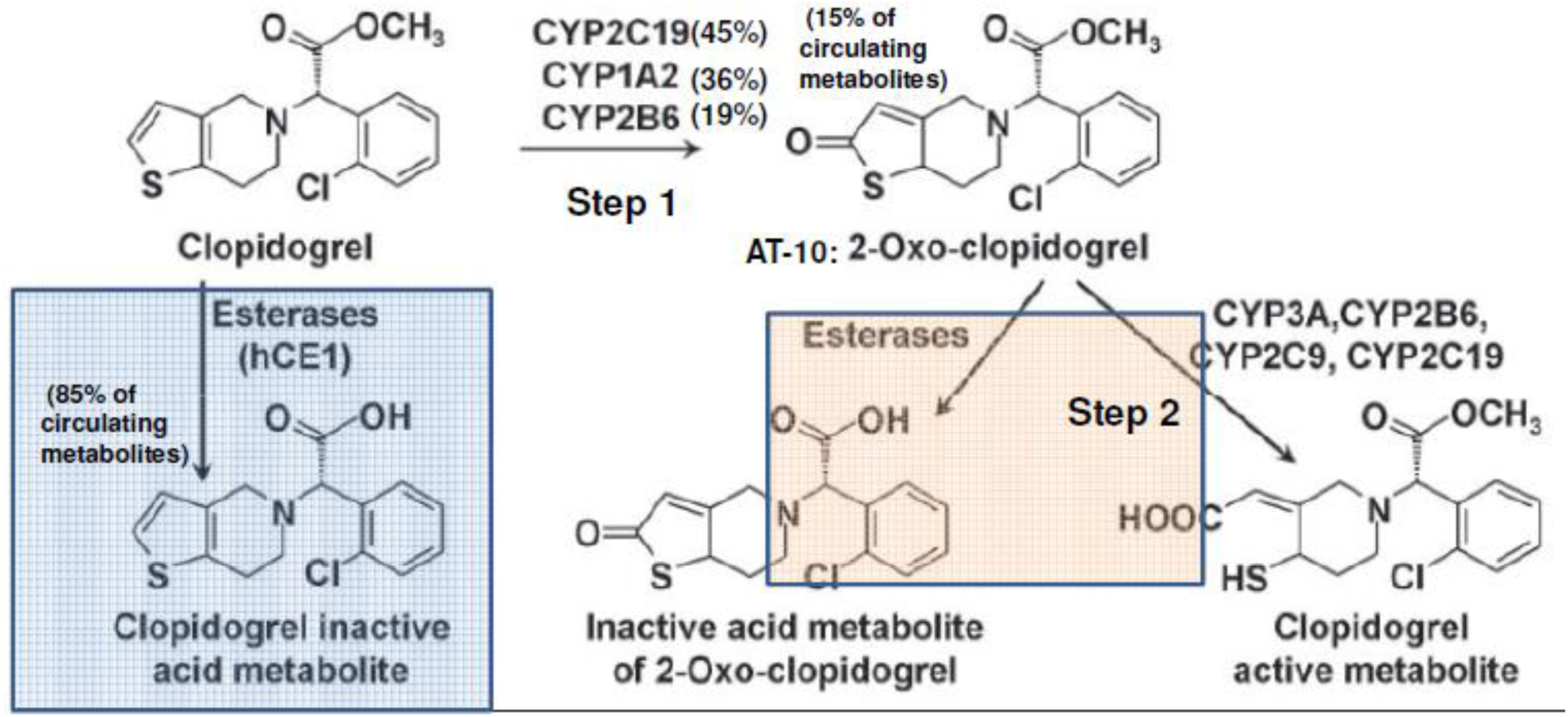
Metabolic Pathway of Clopidogrel and AT-10 (2-Oxo-Clopidogrel)

The response to AT-10 may not be influenced by genetic CYP2C19 polymorphisms to the extent of its influence in Clopidogrel, thus eliminating the need for characterization of Clopidogrel responsiveness and thereby maintaining effectiveness in all patients, including those patients who are identified as CYP2C19 poor and intermediate metabolizers. Prior to this Indian Phase 1 study, one proof of concept clinical study was performed with an objective of evaluating and establishing the dose-response relationship of single and multiple doses of AT-10 ranging from 7.5 to 40 mg in healthy human subjects. In that proof of concept study, AT-10, 40-mg dose was found to be comparable to the Clopidogrel 300 mg dose with regard to %IPA at 6 hr following the single loading dose. Hence, AT-10 40 mg was selected as the loading dose. Since the maintenance dose of Clopidogrel (75 mg) is ¼th of the loading dose of Clopidogrel (300 mg), AT-10 10-mg (¼th of the loading dose of AT-10 40-mg) was selected as the maintenance dose to administer in this Phase 1 Indian study. AT-10 was found to be safe and well tolerated in humans at all doses tested in that proof of concept study. So, in light of promising data obtained from nonclinical and earlier conducted proof of concept clinical study, we first time evaluated safety, tolerability, pharmacodynamics and pharmacokinetics of AT-10 in healthy Indian subjects in comparison with standard dosage regimen of Clopidogrel. It is important to note that AT-10 is an intermediate metabolite in the metabolic pathway of Clopidogrel to the active metabolite. Therefore, this study does not represent the first exposure of AT-10 in humans. It does, however, represent the direct administration of AT-10 to humans.

## METHODS

### Study Design

This was an open label, unicentric, randomized, two-period, two-sequence, crossover, comparative, single and multiple-dose, Pharmacokinetic (PK) and Pharmacodynamic (PD) study conducted in healthy human subjects. Two CYP2C19 genotype based groups were identified i.e., PM (PM) and EMs (EM) with 20 subjects in each group (N=40). Each metabolizer group was further randomized in 1:1 ratio to receive either Clopidogrel (Plavix, Sanofi Aventis, USA) or AT-10 (Ipca Laboratories Limited, India) in 2 sequences. Sequence 1 includes administration of AT-10 on 1^st^ period and Clopidogrel on 2^nd^ period while sequence 2 was the reversal of sequence 1. All subjects underwent a 2 period treatment regimen; each period treatment lasted for 6 days. A study period included administration of loading dose (LD) of Clopidogrel 300 mg or AT-10, 40 mg on Day 1 followed by once daily maintenance dose (MD) of either Clopidogrel 75 mg or AT-10, 10 mg from day 2 to day 6. Then the subjects were crossed over to receive the alternate therapy for the 2^nd^ period. A wash out period of at least 14 days was maintained between the study periods.

On day 1, all subjects fasted for at least 10 hours prior dosing and at least 4 hours post dose in each period. Water was restricted prior to 1 hour of study drug administration and 1 hour post dosing in each period. All subjects were housed in the clinical facility from at least 11 hours prior dosing until at least 24 hours post dosing after administration of last MD in each period.

### Study Population

Healthy Indian adult male and female subjects of age between 18 to 45 years with body mass index (BMI) of 18.50 to 29.90 kg/m^2^ were enrolled in the study. Prior enrollment, we conducted CYP2C19 genotyping for categorizing the subjects as EMs (EM; genotype *1/*1) or PMs (PM; genotype *2/*2). In order to confirm and cross verify the detected genotypes, the blood samples of all screened subjects were assessed independently at two different laboratories. Subjects with matching alleles only at both the testing laboratories were considered for enrollment in the study provided that such subjects meet the study enrollment criteria. Polymerase Chain Reaction (PCR) followed by DNA sequence analysis technology was used for genotyping test. Subjects with no evidence of underlying disease, medical history, clinical examination and laboratory investigations performed during the pre-study screening within 28 days prior to commencement of the study were included.

Subjects with known allergy to Clopidogrel, AT-10 or any component of the formulation and to any other related class of drug; history or presence of significant cardiovascular, respiratory, hepatic, renal, hematological, gastrointestinal, endocrine, immunologic, dermatologic, musculoskeletal, neurological or psychiatric disease; history or presence of significant alcohol dependence (abuse) or drug abuse within the past 1 year; history of chronic smoking or chronic consumption of tobacco products; history of difficulty in donating blood and subjects with known increased risks of bleeding were excluded from the study. Also, Subjects who had donated blood or loss of blood 50 ml to 100 ml within 30 days or 101 ml to 200 ml within 60 days or > 200 ml within 90 days prior to first dose of study medication were excluded. Subjects who had consumed any medication known to alter hepatic metabolic enzyme activity within 28 days prior to the initial dose of study medication (e.g. strong CYP3A inhibitors or inducers, Omeprazole or other proton pump inhibitors) were excluded. Also, Pregnant or lactating women were not included in the study.

### Ethics

The study was done in accordance with the ethical principles of Declaration of Helsinki and requirements of Good Clinical Practice. It was approved by Sangini Hospital Ethics Committee and Indian Regulatory Authority [Central Drug Standard Control Organization (CDSCO)]. It was registered on Clinical Trials Registry India with registration number CTRI/2021/03/032206 [Registered on: 23/03/2021]. Written informed consent was obtained from all the participants’ prior participation in the study. No study related procedure was undertaken prior to obtaining written informed consent from subject.

### Safety Assessment

Safety assessments were performed by the investigator at screening and continued throughout the course of the study. Safety evaluations involved assessment of adverse events, coagulation parameters (PT and PTT), pre-dose and post-dose vital signs, ECGs, clinical laboratory testing (hematology, blood chemistry, and urinalysis), and general physical and clinical examinations. Coagulation parameters were also assessed at screening, at housing during each period, at 8 hrs post-dose on each dosing day and at exit or early termination.

### Pharmacodynamic Assessment

Pharmacodynamic (PD) assessment i.e. inhibition of platelet aggregation was performed by using ADP-induced platelet aggregation from whole blood treated with sodium citrate (3.2%) as an anti-coagulant. The whole blood impedance aggregometry (CHRONO-LOG^®^ 592, Pennsylvania, United States) was used for assessing ADP-induced platelet aggregation. It uses electrical impedance to measure platelet aggregates in response to agonist addition in whole blood. As a result, electrical resistance between the wires increase and this change in impedance is recorded continuously in AGGRO/LINK^®^-WBA software. This change in electrical impedance is directly proportional to the extent of platelet aggregation and is indicated on the digital display in ohms (unit) after six minutes. The inhibition of platelet aggregation was further calculated by using the formula: Platelet Inhibition (%IPA) = (1 – Residual Impedance (amplitude in ohm) / Baseline Impendence (amplitude in ohm)) × 100. The PD samples were analyzed at the clinical site only. Aggregometry was carried-out after stimulation of collected blood sample with 20 μM of ADP. As compared to conventional Light Transmission Aggregometry (LTA), the impedance aggregometry assesses platelet function under more physiological conditions as it is performed in whole blood, thus enabling other blood elements to influence platelet aggregation. Also, it takes place on a solid surface resembling the physiological process of platelet adhesion and aggregation. The principle of impedance method is similar to that of LTA except that it is done in whole blood, thus obviating the need for preparation of a platelet rich and poor plasma which may add variability to the test results.

For pharmacodynamic assessment, blood samples were collected into pre-labeled Citrate Tubes (3.2%). A total of 4 blood samples (2.7 mL each) were collected, prior administration of drug and at 30 minutes, 2 and 6 hours after the study drug administration on each dosing day. Additionally, one more blood sample was collected on day 7 after 24 hours of last MD administration in each period.

### Pharmacokinetic Assessment

In each period, blood samples (5 mL each) were collected in pre-labelled Na-Citrate vacutainers at pre-dose and at 0.25, 0.5, 0.75, 1, 1.5, 2, 3, 4, 6, 8, 12 and 24 h after study drug administration on Days 1 and 6 for assessment of PK parameters. In between on Day 3, 4 and 5 of each period, pre-dose sample was also collected.

Plasma concentrations of derivatized active metabolite H4 (measured as MP-H4) were determined using a validated Liquid Chromatography-Mass Spectrometry (LCMS/MS) method. The PK analysis was performed using a model independent software program (WinNonlin Professional version 8.3). Maximum concentration (C_max_), and the time to reach Cmax i.e., T_max_ were obtained from plasma concentration time curves. Then elimination rate constant (k), elimination half life (t^1/2^), Clearance (Cl), Volume of distribution (V_d_) and Area under the curve (AUC) were calculated following non-compartmental analysis of plasma time course data for active metabolite MP-H4.

### Statistical Analysis

For PD analyses, %IPA generated by Aggregometry at different time points among the groups were assessed. Maximum %IPA for each group was compared using Analyses of variance (ANOVA) with sequence, treatment, period and subject (sequence) as fixed effects. For the comparison between types of metabolizers, the ANOVA model with types of metabolizers as fixed effect was used. Least-squares means (LSMs) with 95% CI was calculated by ANOVA for each comparison.

To estimate the PK parameters, statistical comparison of the overall PK (Day 1 and Day 6), single-dose and multiple-dose PK parameters of peak (Cmax) and total (AUCt, AUCinf) exposure of MP-H4 were made between the AT-10 and Clopidogrel dose groups; between and within metabolizer groups. Geometric means were calculated for Cmax, AUCt and AUCinf. ANOVA was performed on the ln-transformed PK parameters Cmax, AUCt and AUCinf. Geometric means and its 90% CI were given for all parameters in each phenotype.

Data were presented using descriptive statistics i.e. n (%) for categorical variables and mean, SD, minimum and maximum for continuous variables. according to the regulatory guidelines. Significance was calculated using P values; p <0.05 was considered statistically significant. All the statistical analyses were performed using SAS version 9.4.

## RESULTS

### Subject enrollment and Demographics characteristics

A total of 40 subjects were enrolled in the study. Two CYP2C19 genotype based groups were identified i.e., PM and EMs with 20 subjects in each group. Out of 40 enrolled subjects, 37 subjects had completed the study. Remaining 3 subjects were discontinued from the study due to adverse event (n=2) and consent withdrawal (n=1). The two treatment groups were well balanced with regards to all baseline characteristics. Demographic characteristics of these subjects are given in **Table 1**.

**Table 1:** Demographic and baseline characteristics of the enrolled subjects.

| Characteristic | Treatment Sequence |  | Overall<br>(N=40) | P- Value |
| --- | --- | --- | --- | --- |
|  | Sequence AB<br>(N=20) | Sequence BA<br>(N=20) |  |  |
| Gender, n (%) |  |  |  | 0.9999 |
| Female | 6 (30.0) | 6 (30.0) | 12 (30.0) |  |
| Male | 14 (70.0) | 14 (70.0) | 28 (70.0) |  |
| Age (years), Mean ± SD | 32.6 ± 6.36 | 34.9 ± 5.21 | 33.7 ± 5.85 | 0.2185 |
| BMI (Kg/m<sup>2</sup>), Mean ± SD | 23.41 ± 3.47 | 24.51 ± 2.67 | 23.96 ± 3.11 | 0.2691 |
| Height (cm), Mean ± SD | 166.2 ± 10.20 | 162.0 ± 7.23 | 164.1 ± 8.99 | 0.1413 |
| Weight (Kg), Mean ± SD | 64.91 ± 12.67 | 64.08 ± 9.07 | 64.50 ± 10.89 | 0.8119 |
| Race, n (%) |  |  |  |  |
| Asian | 20 (100) | 20 (100) | 40 (100) | 0.9999 |
| Menopause Status, n (%) |  |  |  |  |
| Not Menopausal | 6 (30.0) | 6 (30.0) | 12 (30.0) | 0.9999 |
| Menstrual Cycle, n (%) |  |  |  |  |
| Regular and Clinically Normal | 6 (30.0) | 6 (30.0) | 12 (30.0) | 0.9999 |
| N: The number of subjects in the safety population for each sequence or the safety population for the overall; n: The number of subjects in the specific category. %: calculated using the number of subjects in the safety population for each sequence, or the safety population for the overall, as denominator (n/N*100)<br>Age, Height, weight and BMI at Pre-Screening visit are considered.<br>Treatment A : AT-10 (2-oxo-Clopidogrel bisulfate) Tablets 40 mg/10mg<br>Treatment B : Plavix® (Clopidogrel bisulfate) Tablets 300 mg/75mg |  |  |  |  |

### Pharmacodynamic Results

The ADP induced platelet aggregation was assessed for change from baseline in each treatment group at each time point assessed on each dosing day for PD analysis. The inhibition of ADP-induced platelet aggregation occurred rapidly, reaches peak at around 6 hr and inhibition was sustained until 24hr post dose. The mean %IPA over 6 days of drug administration (loading and maintenance dose) is shown in **Figure 2** for both AT-10 and Clopidogrel.

**Fig.2:**
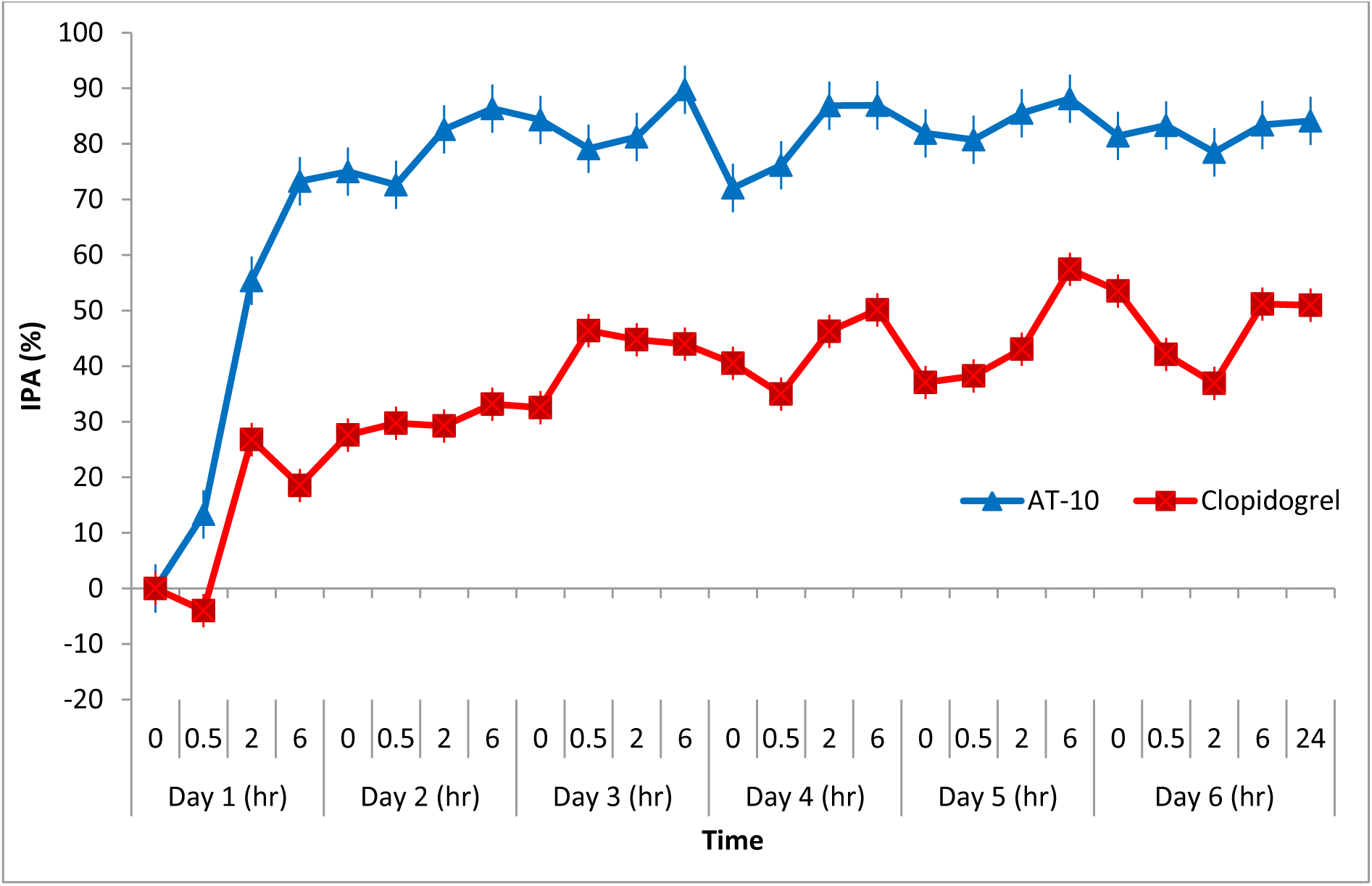
Overall %IPA of AT-10 vs. Clopidogrel.

The mean %IPA between PMs and EMs in AT-10 group showed that, %IPA at Post-dose 6 hr on day 6 (MD) obtained from the AT-10 in EMs group appeared to be higher than the AT-10 treatments in PMs group. However, the difference between the AT-10 treatments in EMs versus AT-10 treatments in PMs was found to be statistically insignificant at post dose 6 hr LD and post dose 24 hr after day 6 (MD). The mean %IPA was found to be comparable in poor and EMs in AT-10 group. It means AT-10 produces equivalent PD effect in both extensive and PMs and the antiplatelet effect of AT-10 is not affected by the genotyping metabolizing status of the volunteers. Although %IPA post 6 hr on day 6 was significant, it was more than 70% in PMs. That should be enough to be efficient. This indicates that the PD response to AT-10 may not be influenced by CYP2C19 polymorphisms in PMs **(Table 2).**

**Table 2:** Mean %IPA between Poor and Extensive Metabolizers in AT10 and Clopidogrel.

| Time point | Extensive Metabolizers | Poor Metabolizers | 95% CI of LS mean difference | P value |
| --- | --- | --- | --- | --- |
| **AT-10** |  |  |  |  |
| Post-dose 6hr (LD) | 80.694 ± 27.5592 | 65.905 ± 30.2122 | 14.79 (-3.72 - 33.30) | 0.1141 |
| Post-dose 6hr on Day 6 (MD) | 94.269 ± 12.9226 | 72.558 ± 40.9339 | 21.71 (1.74 - 41.68) | 0.0340 |
| Post-dose 24hr after Day 6 (MD) | 88.995 ± 21.1644 | 79.032 ± 34.8512 | 9.96 (-9.16 - 29.09) | 0.2975 |
| **Clopidogrel** |  |  |  |  |
| Post-dose 6hr (LD) | 37.140 ± 50.8991 | -1.168 ± 46.1272 | 38.31 (4.84 - 71.78) | 0.0262 |
| Post-dose 6hr on Day 6 (MD) | 71.551 ± 29.9885 | 30.833 ± 42.7628 | 40.72 (15.70 - 65.74) | 0.0022 |
| Post-dose 24hr after Day 6 (MD) | 65.306 ± 58.5336 | 36.644 ± 46.0291 | 28.66 (-5.98 - 63.31) | 0.1020 |
| Data Presented as Mean± SD; LD- Loading Dose; MD-Maintenance Dose |  |  |  |  |

On the contrary, following loading and maintenance dose post 6 hr on Day 6 in Clopidogrel treatment group the mean %IPA was found to be significantly higher for EMs as compared to PMs. In PMs, %IPA observed was half of that observed in EM. This indicates that the PD response to Clopidogrel following loading and maintenance dose is influenced by CYP2C19 polymorphisms resulting in less PD effect in PMs than that in EMs which is consistence with data reported for Clopidogrel in various studies^25,26^. **(Table 2)**

Further, when we compared the mean %IPA between AT-10 PMs and Clopidogrel EMs, the %IPA at Post-dose 6hr (LD) obtained from the AT −10 treatment in PM group was statistically significant with that of Clopidogrel treatments in EMs group. However, %IPA at Post-dose 6 hr on Day 6 (MD) and Post-dose 24 hr after Day 6 obtained from the AT-10 treatment in PM group appears to be statistically insignificant than the Clopidogrel treatments in EMs group. This indicates that the response to AT-10 is not influenced by CYP2C19 polymorphisms, hence patients including those who are CYP2C19 Poor Metabolizers, may receive clinical benefits from AT-10 treatment **(Table 3).**

**Table 3:**
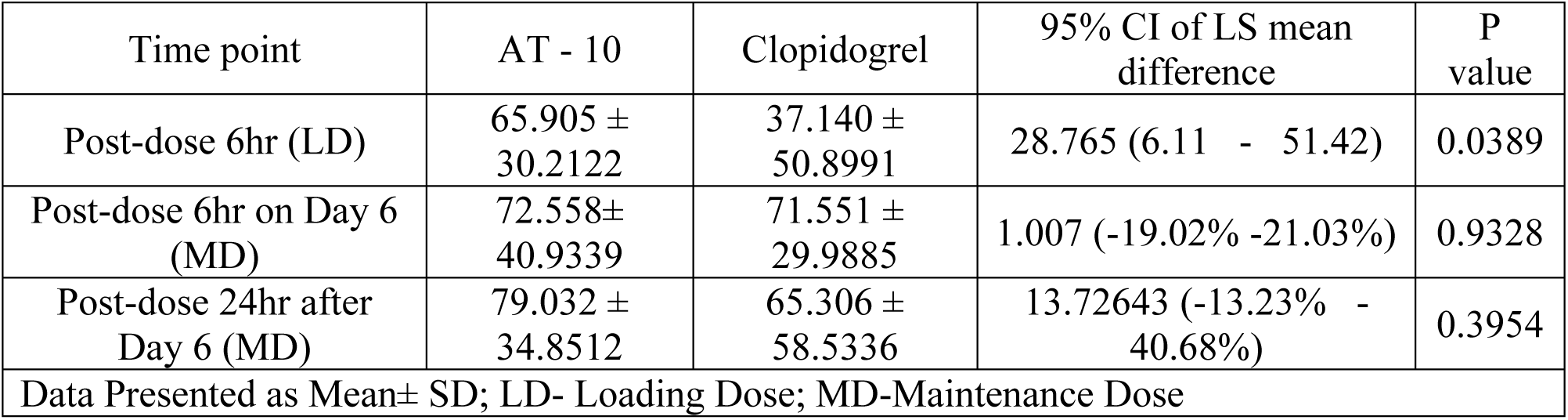
Statistical Analysis of Mean %IPA between AT-10 Poor Metabolizers and Clopidogrel Extensive Metabolizers.

Moreover, the overall result of PD parameters showed that, mean %IPA between AT-10 and Clopidogrel in all subjects at Post-dose 6hr (LD), Post-dose 6 hr on Day 6 and Post-dose 24 hr after Day 6 obtained from the AT-10 group was significantly higher than the Clopidogrel group. Similarly, following loading and maintenance dose, the mean %IPA for AT-10 treatment group was significantly higher than that observed with Clopidogrel treatment group in both poor and EMs **(Figure 2**).

### Pharmacokinetic Results

AT-10 is a Clopidogrel derivative that is metabolically converted to produce the active metabolite (MP-H4) of the approved listed drug Plavix (Clopidogrel bisulfate). The PK parameters for MP-H4 for all subjects have been presented in **Table 4** after administration of AT-10 and Clopidogrel.

**Table 4:** Pharmacokinetic Parameters of MP-H4 on Day 1 and Day 6 (All subjects)

| Parameters | Day1 (N=37) |  | Day6 (N=37) |  |
| --- | --- | --- | --- | --- |
|  | AT-10 | Clopidogrel | AT-10 | Clopidogrel |
| Cmax(ng/mL) | 9.56 ± 7.01<br>(73.33%) | 4.31 ± 4.06<br>(94.30%) | 3.01 ± 1.58<br>(52.35%) | 1.22 ± 0.64<br>(52.51%) |
| #Tmax (hr) | 0.50<br>(0.25 - 2.00) | 0.75<br>(0.25 - 2.00) | 0.50<br>(0.00 - 4.00) | 0.50<br>(0.50 - 1.00) |
| AUCt<br>(hr* ng/mL) | 14.23 ± 4.82<br>(33.84%) | 4.80 ± 2.43<br>(50.57%) | 3.42 ± 1.03<br>(30.26%) | 1.23 ± 0.62<br>(50.52%) |
| AUCinf<br>( hr*ng/mL ) | 15.36 ± 4.41<br>(28.68%) | 5.34 ± 2.36<br>(44.12%) | 3.81 ± 1.12<br>(29.41%) | 1.44 ± 0.63<br>(43.75%) |
| T½ ( hr ) | 3.33 ± 1.85<br>(55.50%) | 0.91 ± 0.43<br>(47.30%) | 1.76 ± 2.29<br>(130.15%) | 0.71 ± 0.55<br>(77.74%) |
| Kel ( 1/hr ) | 0.27 ± 0.13<br>(49.06%) | 0.87 ± 0.27<br>(31.08%) | 0.69 ± 0.40<br>(57.85%) | 1.27 ± 0.50<br>(39.56%) |
| Kel_lower | 3.16 ± 1.77<br>(56.00%) | 1.54 ± 0.67<br>(43.42%) | 1.84 ± 1.03<br>(56.24%) | 0.98 ± 0.23<br>(23.01%) |
| Kel_upper | 11.03 ± 4.51<br>(40.92%) | 4.21 ± 1.14<br>(27.04%) | 4.57 ± 1.58<br>(34.50%) | 2.25 ± 0.62<br>(27.49%) |
| Vd/F (mL) | 2796782.26 ±<br>720716.15<br>(25.77%) | 68069565.39 ±<br>31035114.97<br>(45.59%) | 2914055.03 ±<br>1169552.58<br>(40.13%) | 61932416.51 ±<br>26502532.12<br>(42.79%) |
| Cl/F (mL/hr) | 13282418.32 ±<br>7752256.80<br>(58.36%) | 84001569.64 ±<br>42699043.41<br>(50.83%) | 6254732.88 ±<br>6181797.33<br>(98.83%) | 63098668.98 ±<br>57845349.30<br>(91.67%) |
| Data Presented as Mean ± SD (CV %) |  |  |  |  |
| #For T<sub>max</sub> median (min – max) |  |  |  |  |

Analysis of MP-H4 between AT-10 and Clopidogrel in all subjects at Day 6 showed that, Cmax (AT-10: Clopidogrel; 2.68 ng/mL Vs 1.07 ng/mL p = 0.0001) and AUCt (AT-10: Clopidogrel; 3.27 hr*ng/mL Vs 1.09 hr*ng/mL p = 0.0000) of pharmacologically active metabolite MP-H4 are significantly higher in AT-10 treatment than Clopidogrel indicates that AT-10 at 10mg gives greater exposure of MP-H4 metabolite than Clopidogrel 75mg. MP-H4 metabolite is responsible for pharmacological and therapeutic action; hence pharmacodynamic effect (anti-platelet effect) expected at steady state with AT-10 treatment can be greater than Clopidogrel treatment regardless of genotyping status. The mean plasma concentration curve after administration of loading and maintenance dose of AT-10 and Clopidogrel is shown in **Figure 3**.

**Fig 3:**
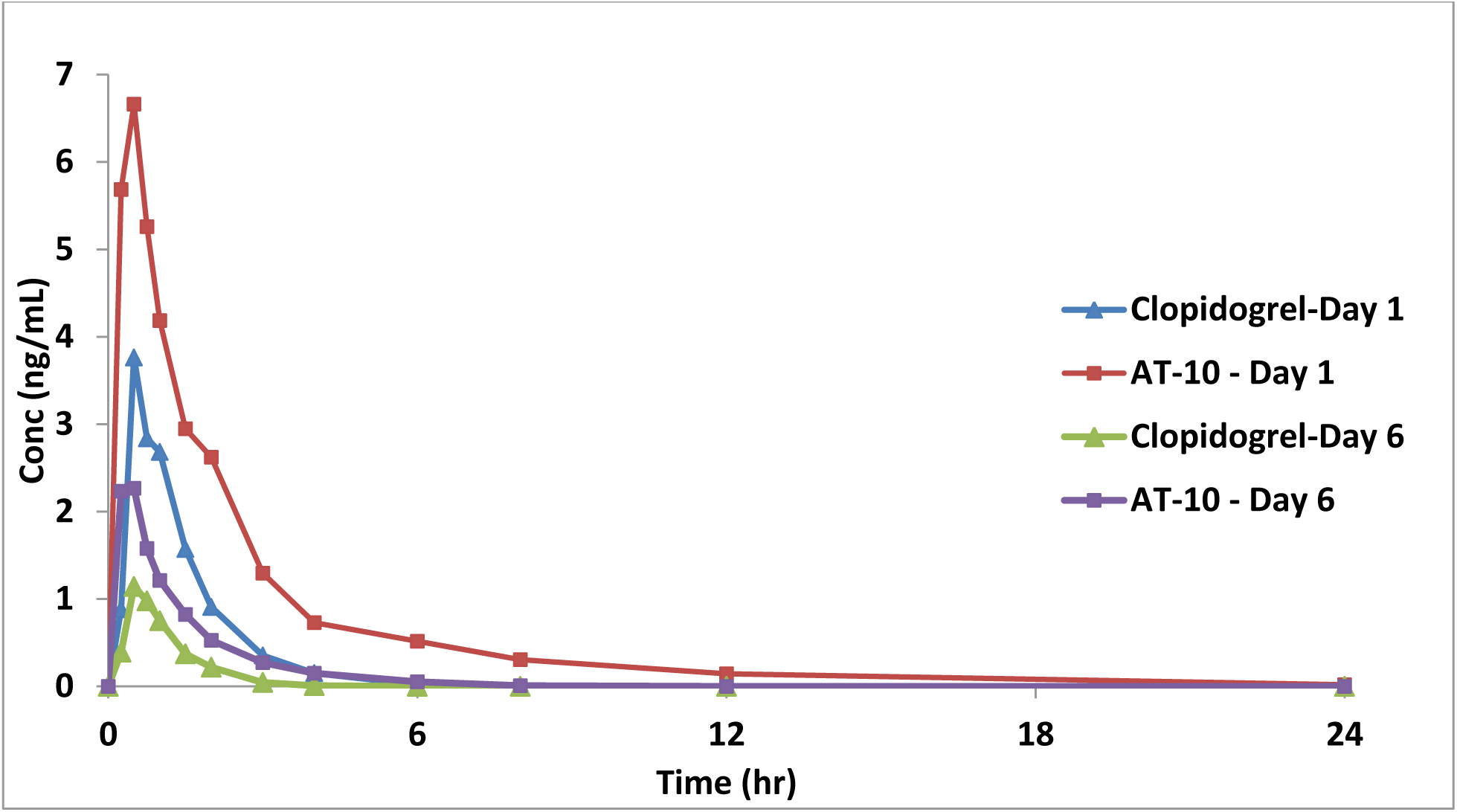
MP-H4 of AT-10 and Clopidogrel on Day 1 and Day 6 (Overall subjects)

Overall, there is a statistically significant difference observed between AT-10 and Clopidogrel treatment for assessed pharmacokinetic parameters. This indicates that 40mg LD and 10mg MD of AT-10 gives greater exposure of H4 (measured as MP-H4) metabolite than Clopidogrel 300 mg loading and 75mg maintenance dose.

### Safety assessment

All enrolled subjects were eligible for safety analysis. Treatment emergent adverse events (TEAE) occurred during the study are summarized in **Table 5**. A total of 11 TEAEs were observed in 7 participants where 5 AEs were mild, 1 AE was moderate and 5 AEs were severe. Nine of these events were possibly related and 2 were unlikely related to the study drug. No serious adverse events were reported during the course of study. Hence, both the treatments were found to be safe and well tolerated in all study population despite the differences in their genotyping status.

**Table 5:** Summary of Treatment Emergent Adverse Events.

| System Organ Class<br>Preferred Term | Treatment A<br>(N = 40) | Treatment B<br>(N = 39) | Overall<br>(N = 40) |
| --- | --- | --- | --- |
| Gastrointestinal Disorders | 2 (0.9) 2 | 3 (1.3) 3 | 5 (12.5) 5 |
| Nausea | 2 (0.9) 2 | 3 (1.3) 3 | 5 (12.5) 5 |
| General Disorders And Administration Site<br>Conditions | 00.0 (00.0) | 1 (0.4) 3 | 1 (2.5) 3 |
| Pain | 00.0 (00.0) | 1 (0.4) 1 | 1 (2.5) 1 |
| Pyrexia | 00.0 (00.0) | 1 (0.4) 1 | 1 (2.5) 1 |
| Swelling | 00.0 (00.0) | 1 (0.4) 1 | 1 (2.5) 1 |
| Psychiatric Disorders | 1 (0.4) 1 | 00.0 (00.0) | 1 (2.5) 1 |
| Anxiety | 1 (0.4) 1 | 00.0 (00.0) | 1 (2.5) 1 |
| Respiratory, Thoracic And Mediastinal Disorders | 1 (0.4) 1 | 00.0 (00.0) | 1 (2.5) 1 |
| Cough | 1 (0.4) 1 | 00.0 (00.0) | 1 (2.5) 1 |
| Skin And Subcutaneous Tissue Disorders | 00.0 (00.0) | 1 (0.4) 1 | 1 (2.5) 1 |
| Erythema | 00.0 (00.0) | 1 (0.4) 1 | 1 (2.5) 1 |
| MedDRA version 24.1. |  |  |  |
| Data presented as n (%) E |  |  |  |
| E: Number of events; N: Number of subjects dosed with each treatment; n: Number of subjects with adverse event with particular category; %: Calculated using the number of subjects treated with each treatment as the denominator (n/N*100). |  |  |  |
| Treatment A: AT-10 (2-oxo-Clopidogrel bisulfate) Tablets 40 mg/10mg |  |  |  |
| Treatment B: Plavix® (Clopidogrel bisulfate) Tablets 300 mg/75mg |  |  |  |

## DISCUSSION

Despite of newer antiplatelet agents (P2Y12 inhibitors), Clopidogrel is the most widely prescribed thienopyridine with acceptable safety profile; however has major shortcoming associated with CYP2C19 genetic polymorphism. After more than a decade following the introduction of dual antiplatelet therapy as the standard of care in the setting of ACS and PCI^11,27^ significant concerns with the effectiveness of Clopidogrel were raised, mainly non-responsiveness to therapeutic effects resulting from CYP polymorphisms^28,29^.

To overcome the limitations of Clopidogrel associated with CYP2C19 metabolism, Ipca has developed a novel antiplatelet drug AT-10, that is metabolically converted to produce the active metabolite of the approved listed drug Plavix (Clopidogrel bisulfate). A major advantage of the AT-10 is its more efficient metabolism as compared to the two-step process for Clopidogrel involving several CYP450 isoenzymes. This study is the first to evaluate the PD, PK, safety and tolerability of AT-10 in Indian subjects. We have also emphasized the effect of a person’s CYP genotype on pharmacological profiles of AT-10 (test drug) and Clopidogrel (reference drug) as previously published studies have shown impact of CYP2C19 polymorphism on Clopidogrel responsiveness up to a major extent ^16,25,30,31^.

This study met its primary objective and showed that AT-10 is safe and well-tolerated when administered as LD of 40mg followed by 10mg MD for 5 consecutive days. Safety findings from all enrolled subjects did not show any clinically significant trends or abnormalities in liver function tests, clinical chemistry, hematology, and coagulation. Most of the AEs were relatively minor in nature and were considered possibly and unlikely related to study drug treatment. Results from vital signs and ECGs were unremarkable. The overall number of subjects with TEAEs was similar in two treatment groups. No bleeding or other signs of bleeding were reported in the subjects for both the treatments.

Following loading and maintenance dose, the mean %IPA for AT-10 treatment group was significantly higher than that observed with Clopidogrel treatment group in overall study population, poor and EMs. It means AT-10 produces higher platelet inhibition than the Clopidogrel following loading and maintenance doses not only in EMs but also in PMs. Hence AT-10 is likely to benefit all patients irrespective of their genotyping status.

The single and multiple dose pharmacokinetics of active metabolite (H4) have been assessed. From the PK data it was observed that, in overall population for MP-H4 the concentration and extent of exposure (Cmax and AUC) following loading and maintenance dose are significantly higher in AT-10 treatment group than Clopidogrel treatment. Similarly, in both extensive and PMs the concentration and extent of exposure (Cmax and AUC) following loading and maintenance dose are significantly higher in AT-10 treatment group than Clopidogrel treatment for MP-H4. It means the pharmacokinetics of Clopidogrel is influenced by genetic CYP2C19 polymorphisms to a greater extent than its influence in AT-10 treatment group. As per the published literature^32^ a strong correlation between H4 plasma exposure and platelet inhibition exists, demonstrating that individuals with highest exposure to H4 active metabolite have the greatest antiplatelet response. Since H4 metabolite of Clopidogrel is responsible for pharmacological and therapeutic action the pharmacodynamics effect expected with AT-10 treatment may be greater than Clopidogrel treatment in both CYP2C19 EMs and PMs.

The HOST-EXAM (Harmonizing Optimal Strategy for Treatment of Coronary Artery Stenosis-Extended Antiplatelet Monotherapy) Extended study had compared the outcomes of Aspirin monotherapy with those of Clopidogrel monotherapy after PCI^33^. Total of 5438 patients who were maintained on DAPT without clinical events for 12±6 months after percutaneous coronary intervention with drug-eluting stents were enrolled. During an extended follow-up of more than 5 years after randomization, Clopidogrel monotherapy (75mg) compared with Aspirin monotherapy (100mg) was associated with lower rates of the composite net clinical outcome (death, nonfatal myocardial infarction, stroke, readmission attributable to acute coronary syndrome, and bleeding) in patients without clinical events for 12±6 months after percutaneous coronary intervention with drug-eluting stents. 12.8 Vs 16.9% (hazard ratio, 0.74 [95% CI, 0.63-0.86]; P<0.001).

As per the recently published Guideline for the Management of Patients With Chronic Coronary Disease by American Heart Association/American College of Cardiology Joint Committee, in patients with Chronic Coronary Disease (CCD) treated with PCI, DAPT consisting of Aspirin and Clopidogrel for 6 months post PCI followed by single antiplatelet therapy is indicated to reduce MACE and bleeding events^34^.

These findings and clinical practice guidelines again supports the use of Clopidogrel as the preferred antiplatelet agent over Aspirin for the secondary prevention of coronary artery disease. It means Clopidogrel has a place in therapy so as AT-10 as it overcomes the shortcomings of Clopidogrel associated mainly with CYP2C19 polymorphism.

As concluded by Jean-Sébastien Hulot et al^26^ in their pharmacogenetic study, this study results also shows that the CYP2C19*2 mutant allele is a major genetic determinant of the pharmacodynamic response to standard-dose Clopidogrel (75 mg/day) in healthy human subjects. As per Drug Label Information of Plavix^15^, Clopidogrel 75mg per day has shown average platelet inhibition of 40% to 60% at steady state but several clinical trials have shown variability in response to inhibition of platelets and are generally around 30% which was also observed in our study.

It is of due importance that CYP2C19 remains the predictor of variability in response of Clopidogrel and has been clearly evident in previous clinical evaluations^25,26^. Similar observations were also seen in our study as EMs have shown IPA in range of 37% to 71% while participants with CYP2C19 LOF alleles representing PMs have shown very poor inhibition of platelets maximum up to 36% only but for AT-10, the platelet inhibition was consistent despite the disparities in phenotypes of the participants. There was no detrimental impact of CYP2C19 polymorphism on %IPA of AT-10. This indicates that AT-10 can be used in all patients without concerning about the type of metabolizer as compared to selective use of Clopidogrel due to ‘Clopidogrel resistance’. Being a Clopidogrel derivative, AT-10 can provide all benefits of Clopidogrel and at the same time will overcome the shortcomings of Clopidogrel. It can be a safer alternative to newer agents being similar to Clopidogrel. Further it may replace Clopidogrel in the management of cerebrovascular diseases (transient ischemic attack and Stroke) and peripheral vascular diseases.

Since the concentration and extent of exposure (C_max_ and AUC) of active metabolite (H4) and the observed mean %IPA following loading and maintenance dose are significantly higher in AT-10 treatment group than Clopidogrel treatment in both EMs and PMs, there is a need to optimize the dose of AT-10 in target population. Further studies can be planned to assess and find out the PD equivalent dose of AT-10 when administered as a loading and maintenance dose.

### Limitations of Investigation

This was a healthy volunteer study with limited sample size. The interventions in the study were evaluated in poor and extensive metabolizers. Effects of AT-10 need further evaluation in intermediate, rapid and ultra rapid metabolizers whose prevalence is significant in general patient population.

Although inhibition of platelet aggregation is recognized as surrogate marker for incidence of MACE (i.e. higher % inhibition correlates to lesser CV events.), it is still unknown what level of %IPA translates in preventing CV events.

Hence, further larger studies are required to investigate the influence of CYP2C19 functional polymorphisms on the response to AT-10 dose in the clinical setting, and especially on the risk of recurrent thrombotic events during AT-10 treatment including bleeding risk as compared to Clopidogrel therapy.

## CONCLUSION

Based on the results of our study, it could be concluded that AT-10 may emerge as a promising anti-platelet drug and can be further developed in clinical studies for the unmet medical needs of cardiovascular diseases. This might be studied alone or as a part of dual antiplatelet therapy for implementation into future treatment strategies.

## Disclosures

**Dr. Anil Pareek** is the President of Medical Affairs and Clinical Research Department in the pharmaceutical Industry, Ipca Laboratories Limited.

**Mr. Nitin Chandurkar** is the Senior Vice President of Clinical Research and Development in the pharmaceutical Industry, Ipca Laboratories Limited.

**Mr. Vivek Raut** is currently the Deputy General Manager in Clinical Research and Development department at Ipca Laboratories Limited.

**Mr. Kumar Naidu** is Assistant General Manager in Clinical Data Management and Statistical Analysis at Ipca Laboratories Limited.

All authors are employees of Ipca Laboratories Ltd and involved in conceptualization, coordination and execution of the study.

## Data Availability

No, we cannot provide data for privacy reasons.

## Acknowledgements

Authors acknowledge CBCC Global Research LLP, Neuberg Supratech Laboratory & Research Institute Pvt. Ltd., geneOmbio Technologies Pvt. Ltd. and Cliantha Research for their extended support in conducting laboratory assessments. Authors appreciate the help of Dr. Baishakhee Bishoyi, employee of Ipca Laboratories for drafting this manuscript; Mangesh Manchekar and Tushar Jadhao, employees of Ipca Laboratories Limited, for their help in labelling and packaging of clinical trial supplies.

## Abbreviations

ACS: Acute Coronary Syndrome
ADP: Adenosine Diphosphate
AUC: Area under the curve
BMI: Body Mass Index
CAD: Coronary Artery Disease
CDSCO: Central Drug Standard Control Organization
CES-1: Carboxylesterase-1
CVD: Cardiovascular Disease
CYP: Cytochrome P
DES: Drug Eluting Stent
EM: Extensive Metabolizers
IM: Intermediate Metabolizers
LCMS/MS: Liquid Chromatography-Mass Spectrometry
LOF: Loss of Function
LSMs: Least-squares means
PCI: Percutaneous Coronary Intervention
PD: Pharmacodynamic
PK: Pharmacokinetic
PM: Poor Metabolizers
TEAE: Treatment Emergent Adverse Events
USFDA: United States Drug and Food Administration

